# A target product profile for electronic clinical decision support algorithms combined with point-of-care diagnostic test results to support evidence-based decisions during patient consultations by health workers

**DOI:** 10.1101/19008433

**Authors:** Karell G. Pellé, Clotilde Rambaud-Althaus, Valérie D’Acremont, Gretchen Moran, Rangarajan Sampath, Zach Katz, Francis Moussy, Garrett L. Mehl, Sabine Dittrich

**Author notes:** **CORRESPONDING AUTHOR:** Karell G. Pellé, PhD, Foundation for Innovative New Diagnostics, Chemin des Mines 9, 1202 Geneva Switzerland.

## Abstract

Health workers in low-resource settings often lack the support and tools to follow evidence-based clinical recommendations for diagnosing, treating and managing sick patients. Digital technologies, by combining patient health information and point of care diagnostics with evidence-based clinical protocols, can help improve the quality of care, the rational use of resources (humans, diagnostics and medicines) and save patient lives. The development of a target product profile for electronic clinical decision support algorithms (CDSAs) aimed at guiding preventive or curative consultations, and that integrate diagnostic test results will help align developer and implementer processes and specifications with the needs of end-users, in terms of quality, safety, performance and operational functionality. To identify characteristics for a CDSA, experts from academia, research institutions, and industry as well as policy makers with expertise in diagnostic and CDSA development, and implementation in LMICs were convened. Experts discussed the critical characteristics of a draft TPP which was revised and finalised through a Delphi process to facilitate consensus building. Experts were in overwhelming agreement that, given that CDSAs provide patients’ management recommendations, the underlying clinical algorithms should be available in human readable format and evidence-based. Whenever possible, the algorithm output should take into account pre-test disease probabilities, diagnostic likelihood ratios of clinical or laboratory predictors and disease probability thresholds to test and to treat. Validation processes should at a minimum ensure the CDSA are implementing faithfully the evidence-based algorithm they are based on (internal validation through clinical association and analytical validation). Additionally, clinical validation, bringing practice evidence about the impact of the CDSA use on health outcomes, was recognized as a good to have. The CDSAs should be designed to fit within clinic workflows, connectivity challenges and high volume settings. Data collected through the tool should conform to local patient privacy regulations and international data standards.

## INTRODUCTION

Health workers at primary health care (PHC) have the greatest challenge in ensuring appropriate care for their communities. One way to support health workers is through the provision of clinical guidance in the form of clinical practice guidelines or clinical decision support algorithms (CDSA). Clinical practice guidelines are a set of recommendations on diagnostic and treatment modalities based on systematic review of evidence and assessment of the benefits and harms for the patients. The World Health Organisation (WHO) developed several simple clinical decision guidelines to support health workers in low- and middle-income countries (LMICs) in the evaluation and management of clinical problems, with the ultimate goal of improving quality of care[1]. This includes the Integrated Management of Childhood Illness (IMCI) designed in the 1990s for common diseases affecting children younger than 5 years of age[2], which was later adapted for use at the community as the integrated Community Case Management guideline (iCCM)[3]. Other guidelines have been developed and either take an integrated management approach similar to IMCI, such as the Integrated Management of Adolescent and Adult Illness (IMAI[4]), or a disease-specific approach (i.e. WHO’s dengue guidelines for diagnosis, treatment, prevention and control[5]).

While the global health community awaits updated and more comprehensive clinical algorithms, the Fourth Industrial Revolution[6] is seeing developing economies “leapfrogging”[7] mobile technologies across multiple sectors and including for health. As a result, a plethora of digital clinical decision support systems (CDSS) have been implemented in LMICs. CDSS are defined as “digitised job aids that combine individual’s health information with the health worker’s knowledge and clinical protocols to assists health workers in making diagnosis and treatment decisions”[8]. It is well recognized that adherence to standardized clinical practice guidelines is a measurement of quality of care[9,10], and that improving adherence to evidence-based guidelines is associated with better health outcomes[11]. By digitalizing clinical guidelines such as IMCI and iCCM, into electronic CDSAs in its simplest form, a decision tree, several groups have shown significant increase in health worker adherence to guidelines compared to routine care (paper-based IMCI)[12, 13,14].

However, based on IMCI/iCCM, today these CDSAs carry the same limitations than the main guidelines, and still lack point of care (POC) diagnostic tests that could improve the accuracy of diagnoses and treatment recommendations. The only diagnostic test widely used with IMCI/iCCM guidelines is a malaria rapid diagnostic test (mRDT) and while this inclusion was important in the context of “test-and-treat”[15], it is limited to ruling in or out one pathogen alone. Acute febrile illness can be caused by bacterial or viral respiratory infections, or seasonal, geographically localised pathogens, like dengue. Hence, the inclusion of additional simple wet and digital diagnostics into adapted algorithms can reduce antimicrobial over-prescriptions as well as patient outcome[16, 17]. With the growing development of CDSAs, especially related to the management of sick children in LMICs (**Supplemental File 1**), it is important to ensure that new tools meet the needs of the end-user and take into consideration the emerging landscape of diagnostic tools that might be included in new algorithms forming a toolkit consisting of CDSA and diagnostics.

The aim of this work was to define a target product profile (TPP) to inform developers and implementers alike of the requirements for CDSAs aimed at guiding health workers during a preventive or curative consultation with any type of patient, and that have the ability to incorporate diagnostic test results, especially when diagnostics tests have been implemented in pre-existing guidelines.

## 1. METHODS

### 1.1. Draft Target Product Profile

A draft TPP (version 0) for CDSAs combined with POC test results was developed (by: KP, SD, RS, ZK, GLM, FM) based on standard procedure at FIND and WHO, where characteristics are defined as “minimal” and “optimal” criteria (**Supplemental File 2**). “Minimal” is used to refer to the lowest acceptable output for a characteristic and “optimal” for the ideal target of a characteristic. Comments are used to explain or provide examples for the characteristics. As this TPP is for a multi-component toolkit, CDSA and POC diagnostics, the structure of the TPP was adapted to cover the different components and for the purpose of clarity. These sections included: Scope General, Scope Toolkit, Electronic Clinical Decision Algorithm, Point of Care Tools, App, Data and Procurement.

### 1.2. Expert Meeting

To discuss selected aspects of the draft TPP (algorithm validation, performance, machine learning, diagnostic data, disease prediction, clinical workflow and application functionality) (**Supplemental File 2**), a meeting on this subject was convened by FIND and WHO involving 39 experts from academic institutions, industry, private and public sectors from 11 countries[18]. Experts were selected based on their experience in relevant areas of work directly related to digital health (clinical decision support or other digital tools targeting resource-challenges settings) and/or diagnostics (wet and digital). Following the expert meeting the TPP was updated based on expert feedback (version 1.0) (**Supplemental File 3**).

### 1.3. Delphi Process

To facilitate consensus building for the TPP, we followed a Delphi-like process where the TPP version 1.0 was reviewed. For this, a three-part online survey was developed. The first part of the survey collected professional information for survey respondents as well as experience with clinical practice/research, diagnostic development, software development, implementation of healthcare programs in LMICs, and health information systems. The second part of the survey allowed for rating of both minimal and optimal criteria for all TPP characteristics. Agreement was scored on a Likert scale from 1-5, where 1 corresponds to fully disagree, 2-mostly disagree, 3-neither agree nor disagree, 4-mostly agree and 5-fully agree. Disagreement with a criterion was based on a rating from 1 to 3 and required a comment/suggestions by the survey respondent. To reach consensus, more than 75% of survey respondents should provide a rating of at least 4 (agreement) on the Likert scale. The third part of the survey provided an opportunity for respondents to share comments/feedback/ideas on the TPP if not covered in the characteristics section. The survey was developed with the online software SurveyGizmo (SurveyGizmo LLC, Boulder CO, USA). Survey analyses were performed using Microsoft Excel 2013 (Microsoft Corporation, Redmond, USA).

#### Patient and public involvement statement

At no point in this work were patients or the public involved in the design of this study or reporting and dissemination of the results.

## RESULTS

### 1. Expert Consensus on the Target Product Profile through a Delphi Process

In the first round, the online survey was sent to 77 experts (including workshop participants) and 28 responded to the survey (response rate, 36.4%), with some respondents submitting a coordinated response for their institution. Twenty-five percent of respondents were from LMICs (7/28), however the majority of respondents, (22/28, 78%), had experience working in Africa and 57% (16/28) in South East Asia. More than 85% of respondents (24/28) had more than 5 years of experience in clinical practice and research, 50% (14/28) in health programme implementation in LMICs, 39% (11/28) in health information systems, 21% (6/28) in software development and 14% (4/28) in diagnostic product development **(Supplemental File 4)**. Most participants work in international organizations (7/28, 25%) and academic institutions (10/28, 35.7%). In the first round, at least 50% agreement was reached for all criteria for all characteristics and feedback was taken into consideration to develop TPP version 1.1 for the second round of the Delphi survey (**Supplemental File 5**). In this new version, the characteristic “Encounter” was removed as several participants expressed it was redundant with the characteristic “Workflow”. Therefore version 1.1 had 40 characteristics.

A second online survey was sent to the 28 respondents from round 1. In this round, 23 participants responded to the survey (response rate, 82%). Most participants were from academia (7/23, 30%), non-profit international organizations (7/23, 30%) and had a similar profile than respondents from round 1 (**Supplemental File 4**). In this round of review, at least 75% of participants agreed with the minimal and optimal criteria for all TPP characteristics. On average, 91% of survey respondents agreed with the minimal and optimal criteria (**Figure 1**) and the TPP was finalized (version 2).

**Figure 1.**
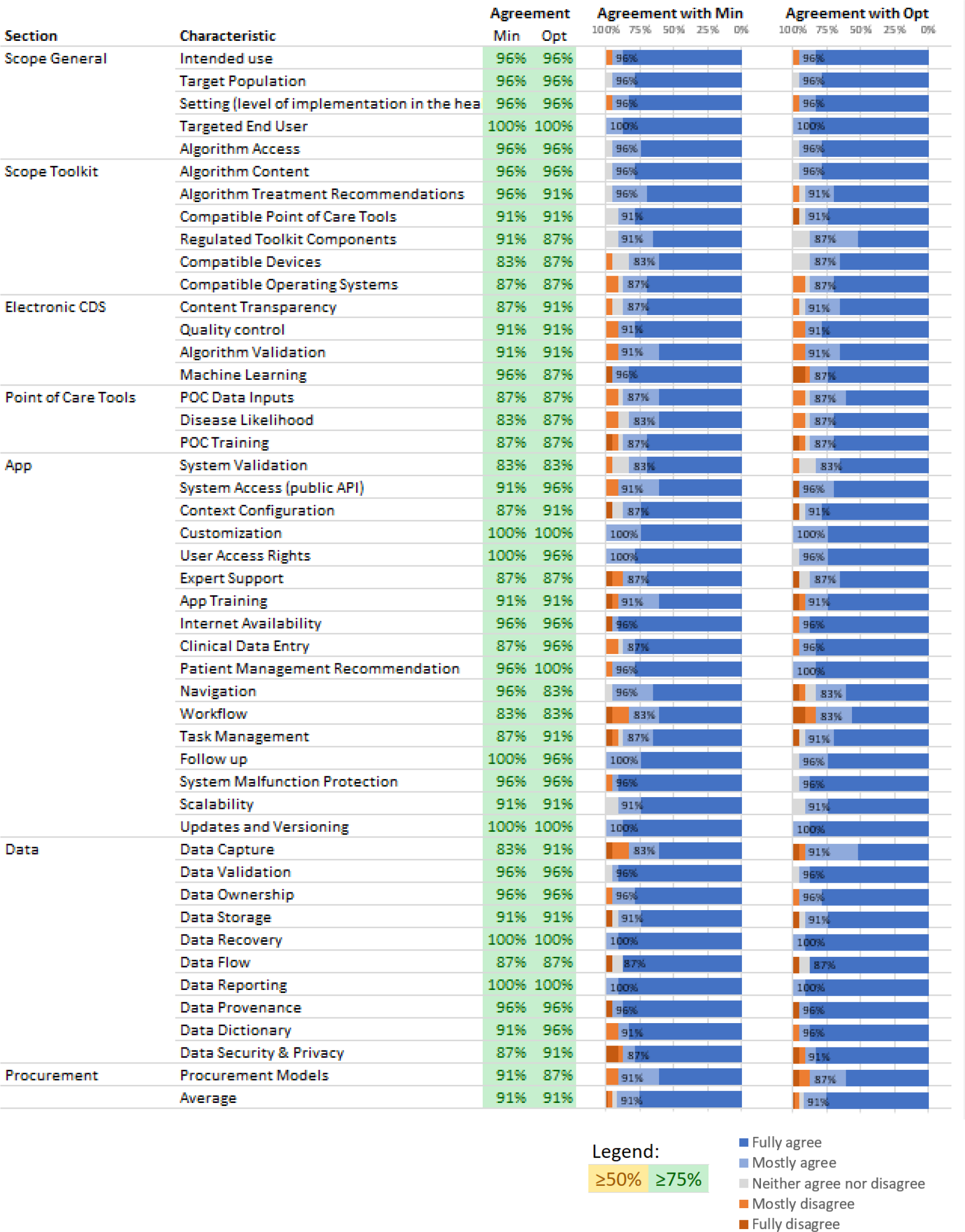
Target product profile second and final survey results.

### 2. Final Target Product Profile

#### Scope General and Scope Toolkit Components

This TPP defines a toolkit composed of CDSAs and POC diagnostics intended to support evidence-based clinical decisions taken by health workers during a preventive or curative consultation, by capturing patient clinical and context-specific data, as well as diagnostic test results to provide recommendations on diagnosis and patient management (including preventive & curative treatment, counseling, etc.). The clinical algorithm integrates POC diagnostic results and is embedded in an App. POC diagnostics in this toolkit should be regulatory approved for use in the setting of implementation.

The experts agreed that the CDSA should first define its target i.e. i) patients (e.g. age group), ii) medical problems (e.g. syndrome or disease), iii) end users (e.g. minimally trained health worker or trained clinician) and iv) level of care (e.g. community or health facility) for which the clinical recommendations are meant and relevant. These specifications should be clearly communicated to the end-user at the beginning of the algorithm. The health worker should immediately be warned if information she or he enters in the system does not fit with these specifications. In cases where an App proposes different algorithms for different use cases (i.e. antenatal care, management of childhood illness), the specifications shall be clearly available for each individual algorithm (**Table 1**).

**Table 1:** Minimal and optimal target product profile characteristics focused on the scope of the toolkit and toolkit component characteristics, as defined by expert consensus process.

| Scope General |  |  |  |
| --- | --- | --- | --- |
| CHARACTERISTIC | MINIMAL REQUIREMENT | OPTIMAL REQUIREMENT | COMMENT |
| Intended Use | The toolkit, composed of an electronic clinical decision support algorithm(s) and point of care diagnostic tests, is intended to increase **evidence-based** treatment decisions by capturing diagnostic test results, patient clinical data (e.g. exposures, signs including vital signs) and context specific data (e.g. disease incidence, seasonality) to provide treatment and care recommendations |  |  |
| Target Population | The algorithm shall define the target population<br>Inclusion and exclusion criteria are used at the encounter to enrol the patient |  | The system can be modular i.e. composed of discrete algorithms such that one can be used for a specific population based on pre-defined criteria |
| Setting (level of implementation in the healthcare system) | Defined by the algorithm |  | The system can be modular i.e. composed of discrete algorithms such that one can be used for a specific healthcare setting based on the infrastructure, workforce knowledge and skills, point of care tools available |
| Targeted End User | Defined by the algorithm |  | The end user shall have the required training/skills to use the App appropriately |
| Scope Toolkit Components |  |  |  |
| Algorithm Access | The electronic clinical decision-support algorithm is accessible through an App downloaded on compatible target devices |  | The App can be a web app, a native app or a hybrid app |
| Algorithm Content | Built on:<br>— well-established clinical evidence based on WHO/international/local clinical care guidelines, peer-reviewed articles (systematic reviews, original clinical research), clinical experience/practice; **and/or**<br>— appropriate clinical validation research* |  | * See FDA's SaMD Clinical Evaluation for guidance[20] |
| Algorithm Treatment Recommendations | Therapeutic recommendations shall be compliant with national treatment guidelines and national EML and support antimicrobial stewardship | Same and evidence based medicine to support optimal treatment recommendations | Recommendations support the appropriate selection, dosage and duration of antimicrobial and any other kind of treatment and management, causing the least harm to the patient |
| Compatible Point of Care Tools | POCs or other relevant medical devices prompted for use by the App shall be locally relevant, i.e. recommended by EDL or relevant national equivalent, or country program | Same, plus emerging diagnostic tools and medical devices relevant to the algorithm and implementation setting |  |
| Regulated Toolkit Components | POC diagnostic tests and medical devices are regulatory approved and compliant with local regulations | Same, and if the software is a medical device, the App shall also have regulatory approval |  |
| Compatible Devices | The App is compatible with any device including:<br>— Smartphones<br>— Tablets<br>— Computers |  | Computers are included as it may be necessary for health facility supervisors to access data collected at the facility level to make informed decisions (i.e. restocking medical supplies) |
| Compatible Operating Systems | OS agnostic | Same as Minimal |  |

#### Clinical Decision Support Algorithms

Regarding the algorithm medical content, the working group agreed that it must be evidence-based. Whether newly developed or adapted from existing guidelines, the algorithm medical content should be developed following international standards for the development or adaptation of evidence-based clinical guidelines, to ensure it implements the best available evidence. This evidence can originate from well-established clinical guidelines such as WHO/international/local clinical care guidelines, or from peer-reviewed articles, clinical practice and/or validation research and the level of evidence and the strength of recommendations in the target context should ideally be communicated. In line with this, recommendations should cause the least harm to the patient and to the community. For example, algorithms related to treatment and management of patient suffering from infectious diseases should also support antimicrobial stewardship to avoid unnecessary side effects for the patient and the development of resistance in the community and at global level. In addition, the algorithm should be validated both analytically and semantically to ensure that the algorithm output is accurate and reproducible, doesn’t deviate from expert content/evidence and that there are no interactions or conflicts in the logic.

Further, important characteristics were content quality, machine learning (ML). With the rise of ML also in this domain of CDSAs to analyse the data captured through algorithms and then for example augment algorithm diagnostic or prognostic accuracy, workshop experts thought it important to address this in the TPP. It was agreed that an algorithm that recommends treatment decisions and management of patients should be at a minimum human interpretable, meaning one should understand how input data are processed into output data. Changes to the algorithm logic based on ML analyses should also be validated (**Table 2**).

**Table 2:** Minimal and optimal target product profile characteristics focused on the electronic clinical decision-support algorithm and point of care tools, as defined by expert consensus process.

| Clinical Decision-Support Algorithm |  |  |  |
| --- | --- | --- | --- |
| CHARACTERISTIC | MINIMAL REQUIREMENT | OPTIMAL REQUIREMENT | COMMENT |
| Content Transparency | The algorithm is “human interpretable”. The healthcare programme and end user can comprehend the algorithm decision-making processes | The healthcare programme and end user have access to underlying evidence and methodology used to develop the algorithm | Human interpretable: a human can understand the choices taken by the model in the decision-making process, i.e. how output variables are generated based on input variables. Visual representations (e.g.. decision trees, Principle Component Analyses, protocol charts, etc.) and performance metrics can be used to support content transparency |
| Quality control | The algorithm has been A) analytically and B) semantically tested:<br>A) Analytical verification: the algorithm output is accurate and reproducible<br>B) Semantical verification: the algorithm doesn’t deviate from expert content/evidence and there are no interactions or conflicts in the logic |  | A) and B) answer the question “did I build the model right?” (See FDA’s SaMD Clinical Evaluation for current guidance[20]) |
| Algorithm Validation | The algorithm has been validated. The level of validation will depend on the CDSS status as a Software as a Medical Device (SaMD)<br>Refer to upcoming rulings from regulatory bodies such as the FDA or the European Commission |  | Answers the question “did I build the right model”? (See FDA’s SaMD Clinical Evaluation for current guidance[20]) |
| Machine Learning | None. The algorithm is static | ML is applied to generate data on the algorithm performance, improve content, inform healthcare system processes, etc.<br>Changes in the algorithm based on ML shall be validated |  |
| **Point Of Care Tool** |  |  |  |
| POC Data Inputs | Any kind of data (qualitative data such as positive/negative/invalid lateral flow test results and quantitative data such as data provided by haemoglobinometers and glucometers) |  |  |
| Disease Likelihood | Based on:<br>— pre-test probability (in the absence of POC or POC performance data); **or**<br>— POC positive/negative likelihood ratio | Based on:<br>— pre-test probability; **and**<br>— POC positive/negative likelihood ratio | The test performance (eg. likelihood ratio) is known and performance data ideally previously determined through independent studies in relevant settings<br>The test brand should ideally also be considered so as to account for changes between manufacturers |
| POC Training | On-site training performed by local authority or implementer |  |  |

#### Point of Care Tools

Experts agreed that the post-test probability or the likelihood of a disease being present, should be assessed based on the pre-test probability—the prevalence of the disease in the corresponding patient population—and the diagnostic positive/negative likelihood ratio—how good a test predicts the presence or absence of disease—of all clinical information i.e. signs, symptoms, and POC diagnostic test results (**Table 2**). Clinical diagnosis often lacks accuracy and therefore diagnostic tests that have the proven added value for patient management should be integrated within algorithms. These tests can be wet diagnostics such as pathogen-specific RDTs or hand-held devices such as haemoglobinometers, and do not exclude digital diagnostics such as digital thermometers.

#### App

An important characteristic in this section is System Validation. Today, there is no standard process to validate and assess performance accuracy of CDSAs. There is also a severe lack of evidence and peer-reviewed publications on both the development and the performance of these tools in LMICs. While some groups have performed randomised controlled trials (RCTs) to measure patient outcome and other parameters that impact patient health, such as the reduction of antibiotic over-prescription[16, 17], others have assessed the performance of their tool by measuring health worker adherence to guidelines[12,13,14]. The level of validation ultimately depends on the type of algorithm. The USFDA has provided some guidance to developers and implementers on the kind of evidence that needs to be provided[19]. The TPP criteria are inspired by this guidance. The minimal criteria guarantee that the App has a CDSA that is based on evidence (clinical association) and that input data is processed correctly (analytical validation). The optimal criteria include clinical validation in addition to clinical association and analytical validation. Through clinical validation, the App has been assessed for the intended outcome (i.e. improved clinical outcome, better quality of care, rational use of resources and reduction of antibiotic over-prescription, etc.) (**Table 3**).

**Table 3:** Minimal and optimal target product profile characteristics focused on App characteristics, as defined by expert consensus process.

| **App** |  |  |  |
| --- | --- | --- | --- |
| **CHARACTERISTIC** | **MINIMAL REQUIREMENT** | **OPTIMAL REQUIREMENT** | **COMMENT** |
| System Validation | The App has been validated in the intended implementation setting. There is evidence demonstrating:<br>— Valid Clinical Association (clinical output based on input data is supported by well-established or novel evidence) **and**<br>— Analytical Validation (input data is processed correctly into expected output data) | The App has been validated in the intended setting. There is evidence demonstrating:<br>— Valid Clinical Association (clinical output based on input data is supported by well-established or novel evidence), **and**<br>— Analytical Validation (input data is processed correctly into expected output data), **and**<br>— Clinical Validation (clinical safety or other meaningful outcome relevant to the intended use) | There is evidence that the system is based on evidence and working to achieve the intended use for the intended setting<br>See FDA's SaMD Clinical Evaluation for current guidance[20] |
| System Access (public API) | Publicly available application programming interface (API) for data access protected by authentication and authorization. At a minimum, technical standards are adhered to | Optimally, HIE/HL7 standards are adhered to |  |
| Context Configuration | — Language: UN official languages<br>— Local time<br>— Local weights and measures | Same as minimal and<br>— Option to customise to local official language<br>— Other country preferences | Language can be a major barrier for the proper use of the tool for patient management and can lead to errors and misinterpretation |
| Customization | Changes to the App, such as updates to the list of medicines and POCs available in the setting, can be made by the healthcare programme. Validation of this change shall be provided. | Same as Minimal |  |
| User Access Rights | Appropriate data access is provided based on specific roles |  | Roles may include data manager (facility supervisor) or data entry person (nurse) |
| Expert Support | None | Built-in access to online/remote expert advice to assist in patient consultation via SMS, audio call, video conferencing |  |
| App Training | On-site training | Same and remote training and/or remote “Train the Trainer” |  |
| Internet Availability | Functions offline (allows for service delivery and key workflows)<br>Allows automatic resynchronisation | Same and trigger alerts on user device when data has not been synchronised for a long time | Internet connection can be very unstable therefore the tool should work in offline mode so as to not disrupt the workflow of the user |
| Clinical Data Entry | Manual entry by the operator | Same, plus automatic upload of digital data (e.g. from biosensors, medical devices) | Optimal: This allows external integration of other App modules, built-in and third party Apps and devices |
| Patient Management Recommendation | Consultation data is summarised and actionable recommendations provided (e.g. treatment, referral, home care or follow up) | Same **and** recommendations are integrated in EMRs and HIS |  |
| Navigation | Sequential: the user follows a strict sequence of data input to reach a final recommendation | Non-sequential: the user can move in any direction through an assessment and change input data to reach a final recommendation |  |
| Workflow requirements to enable time-delayed POC data input | Ability for a user to perform multiple, simultaneous consultations, with pause and resume capability, to allow clinical and laboratory data entry | Same as minimal plus ability to disable simultaneous workflow feature in settings with minimally skilled workers | This is particularly relevant for implementation in settings where testing and clinical consultations are performed in different locations |
| Task Management | Multiple algorithms can be supported simultaneously in one application against a common dataset |  | Can accommodate task-shifting capability i.e. multiple consultations can be opened at a time and patient profiles can be accessed using pre-set user access rights |
| Follow up | None | Ability to retrieve patient information using patient registration information | The optimal implies data recoverability, also covered below in Data Characteristics section |
| System Malfunction Protection | System malfunctions are made clear to the user |  |  |
| Scalability | The App shall allow high transaction volumes with complex workflows to cover primary care workforce at a national scale |  |  |
| Updates and Versioning | Processes are in place to control any App change (including algorithm version updates) and provide the appropriate and correct update to the user |  |  |

In May 2017, a new regulation (EU 2017/745) on medical devices, to which CDSA pertain by definition[20] was adopted by the European Commission, that will be applied starting spring 2020[21]. This regulation includes a reinforcement of the rules on the clinical evidence that should be provided by developers and manufacturers, at least if based in Europe. While waiting for LMICs to have their own regulations, this new regulation will certainly impact development and validation by European developers.

Experts also called for Application Programming Interface (API) to allow other software to ‘talk to’ and interact with the tool through commonly known programming languages. The App should be protected by authorisation and authentication that as a minimum requirement adhere to usual technical standards, and optimally would adhere to HIE (Health Information Exchange) and/or Health Level 7 International[22] medical standards. It was noted that adherence to HL7 can be expensive, hence this was proposed as an optimal rather than a minimal requirement. The App should also be designed to integrate with patient care pathways and health worker workflows with limited disruption. The CDSA accordingly has pause and resume capability to allow time for diagnostic tests to be performed and results entered in the algorithm. POC diagnostic tests such as mRDTs can require up to 20 min for results to be read, depending on the manufacturer.

#### Data

Digital tools have the potential to amass enormous amount of data. With CDSA, patient personal data is captured, stored and potentially used for other operational (i.e. supply chain management) and algorithm development purposes. Many countries have personal data privacy and protection laws in place to protect citizens from the abuse of their personal information. In the European Union this is known as the General Data Protection Regulation[23]. However, there are still many countries around the world that do not have a national eHealth strategy or personal data legislation. This called for the need to address personal data handling with CDSA for security and accountability purposes. Experts agreed that the implementer, who would be considered as the data controller, is responsible for the data and should comply with local data policies and legislations. No specific legislation was mentioned as these differ by country and region. In addition, the historic of the processes that are applied to data as well as the origin of data should be documented (Data Provenance) and the App should function under secure connectivity to avoid loss and corruption of sensitive data, and mitigate cyber-attacks, whether data is at rest or in transmission **(Table 4)**.

**Table 4:** Minimal and optimal target product profile characteristics focused on data characteristics, as defined by expert consensus process.

| **Data** |  |  |  |
| --- | --- | --- | --- |
| **CHARACTERISTIC** | **MINIMAL REQUIREMENT** | **OPTIMAL REQUIREMENT** | **COMMENT** |
| Data Capture | Text, numeric, image, audio, video | Same, and GPS, barcode, biometric |  |
| Data Validation | The system validates data entry to prevent errors that diminish value of the data or the outcome |  |  |
| Data Ownership | Ownership shall be determined by the healthcare programme |  | The healthcare programme is responsible for compliance with any country law, policy and regulation |
| Data Storage | The healthcare programme shall be able to choose the destination of the App's data | Same as Minimal |  |
| Data Recovery | Data can be recovered or the system can be re-established to the desired state in the event of interruption or failure |  |  |
| Data Flow | The flow of data shall be determined by the healthcare programme | Same as Minimal |  |
| Data Reporting | Data export available from all target devices | Pre-built data reporting, analytics and dashboards are available with the App | The level of data manipulation, aggregation and reporting should be sensitive to the device the App is running on i.e. the computer App can be rich in functionality, the mobile App is optimised for data collection and exchange only |
| Data Provenance | Included | Same | Provides origin and processes applied to output data. When data is downloaded or shared, the version of the model is tagged so it is always clear how the data was obtained |
| Data Dictionary | Available, referencing standards used (eg. International Classification of Diseases, Systematized Nomenclature of Medicine) |  | Ensures indicators reported are uniform across different health programs |
| Data Security & Privacy | The App operates under secure connectivity, which meet data protection and regulations of individual countries to avoid loss and corruption of sensitive data, and mitigate cyber-attacks, whether data is at rest or in transmission.<br>Conforms to national privacy laws. Includes processes such as:<br>— Two factor authentication<br>— Authorization/Access Control<br>— De-identified data<br>— Data encryption |  | Encourages GDPR (should no national data security policies exist) to ensure a system that:<br>— preserves data integrity<br>— identifies & mitigates risks<br>— provides relevant parties security processes |

## DISCUSSION

In this work, our aim was to develop a TPP to enable the development of a toolkit that includes CDSAs that are based on evidence and designed to integrate results of diagnostics performed at POC to guide rational clinical decisions. The use of such toolkit would aid in strengthening healthcare particularly at the community and primary care echelons, and help address health workers difficulties to differentiate diseases and syndromes without diagnostic tests and the aid of formalised diagnostic processes that can be provided through algorithms.

In terms of TPP characteristics, “Data” and “Validation” emerged as key topics of discussion. Data generated from CDSA require storage and connectivity and are therefore vulnerable to abuse, loss or unlawful purposes. Although many countries have enacted personal data protection legislation, many fall short on enforcing these and others are still drafting bills. For example, in the African region, in 2017, 17 (out of 54) African countries had put in place comprehensive personal data protection legislation[24]. Several regimes in the region are starting to draft legislation regarding processing and movement of personal data[25], however funding attrition and the lack of regulators cripple the process. That said, there are continent-wide initiatives that promise to set data protection framework such as EU’s GDPR. The African Network of Personal Data Protection Authorities (RAPDP), a cooperation of 8 African countries seeks to draft data protection laws, formulate opinions on specific issues and establish a consultative framework on data protection. There are also regional conventions for data protection and privacy that have been enacted such as the Southern African Development Community (SADC) Model Law[26] for the south and the Act on Personal Data Protection within ECOWAS (Economic Community of West African States) for the west[27]. Unlike their southern and western counterparts, east African nations have not adopted similar regional frameworks, which can set cross-border and data portability limitations. In addition, some countries national laws fall short in setting the required safeguards for data privacy breach and data portability and lack bodies such as Data Protection Authorities to enforce these legislations[24]. This is just the African scenario, but every country and region will have their own set of policies. Therefore, the healthcare programme and data controllers have an important responsibility to manage the programme in compliance with local laws or, where lacking, consult with government authorities on which regulation to abide by to foremost protect patient rights. Regulation of CDSS is also very contentious topic. Groups will have to watch this space, FDA and European Commission regulations, closely to determine whether their tool fits the definition of a Software as a Medical Device (SaMD) and what data they will need to generate for regulatory approval.

In terms of “Validation”, the big question was how to ensure CDSAs are safe for use. Developers and implementers should aim for clinical association, analytical validation and clinical validation. However clinical validation can require significant amount of financial resources and time. As an example, e-POCT was developed in 2014 to incorporate the latest scientific evidence to expand the medical content of IMCI. Its safety and efficacy was determined in a RCT in Tanzanian outpatient clinics[17]. However, in 2019 the tool has yet been implemented at scale or evaluated in non-controlled settings. Efforts like e-POCT to optimise CDSA content and assess its impact on patient health outcomes are needed to grow the evidence for CDSA’s role in improving patient safety. But providing these tools to patient also demands that we put our efforts in studies to measure the effectiveness of CDSA while in the hands of the end-user and in the setting of intended use. Indeed, there may be factors when CDSAs are implemented as part of routine practice that affect the overall effectiveness and impact of the tool. This includes a lack of or poor health worker training, lack of connectivity, software malfunctions, poor design leading to low usability and acceptability of the tool by health workers. Cycles of evaluation and improvement can be sought to perfect these tools in intended settings.

In addition, digital data obtained from CDSA validation can be used in turn to dynamically improve the tool itself faster than development improvements in classic wet diagnostics. CDSA also have the potential to serve as platforms to support the evaluation or development of new content or guidelines. These can be faster adapted to geographies to conform to local policies.

The call for the development of PHC during the declaration of Astana revived the global commitment to provide quality health services to all[28]. Achieving universal health coverage will therefore require that digital health interventions are quality assured and evidence-based. Indeed, the WHO has begun developing frameworks to create a common digital language and synthetizing the evidence around emerging mobile-based digital health technologies through two recent publications[29, 88]. These resources are meant to guide the global health community in assessing digital interventions that will improve quality of care, meaning interventions that are safe, effective, timely, efficient, equitable and people-centred, and this include CDSAs[30].

The management of patients in low-resource settings with complex epidemiology is extremely challenging without clinical algorithms and accurate and portable rapid diagnostic tests. Consequences of suboptimal quality of care can go even beyond the patient’s health to include public health considerations such as the development of antimicrobial resistance due to the over-prescribing of medicines. The aim of the proposed TPP is to support developers and implementers of these toolkits aimed at guiding health workers throughout clinical assessment, to give them confidence in clinical decisions and actions, whether it is sending a patient to a hospital or not, prescribing or not prescribing antibiotics or recommending rehydration at home.

## Data Availability

All data are included in supporting files.

## ACKNOWLEDGEMENTS

We would like to thank all the experts who participated in the FIND/WHO workshop to discuss the first draft of the TPP, as well as Rigveda Kadam, Wallace White and Mo Tobin (FIND) for discussions on TPP development. We would also like to thank the online reviewers who provided feedback to develop the final consensus-driven TPP characteristics.

## FUNDING

This work was funded by the Fondation Botnar and supported by the Global Antimicrobial Resistance Innovation Fund (GAMRIF), a UK aid programme. The funders had no role in the study design, data collection and analysis, or preparation of the manuscript.

## Notes

### Competing Interest Statement

Dr. D'Acremont reports grants from Fondation Botnar, during the conduct of the study; In addition, Dr. D'Acremont has a patent free license on a CDSA called ALMANACH.

### Author Declarations

All relevant ethical guidelines have been followed and any necessary IRB and/or ethics committee approvals have been obtained.

Any clinical trials involved have been registered with an ICMJE-approved registry such as ClinicalTrials.gov and the trial ID is included in the manuscript.

